# Longitudinal Surveillance for SARS-CoV-2 Among Staff in Six Colorado Long-Term Care Facilities: Epidemiologic, Virologic and Sequence Analysis

**DOI:** 10.1101/2020.06.08.20125989

**Authors:** Emily N. Gallichotte, Kendra M. Quicke, Nicole R. Sexton, Emily Fitzmeyer, Michael C. Young, Ashley J. Janich, Karen Dobos, Kristy L Pabilonia, Gregory Gahm, Elizabeth J. Carlton, Gregory D. Ebel, Nicole Ehrhart

## Abstract

**Background:** SARS-CoV-2 emerged in 2019 and has become a major global pathogen. Its emergence is notable due to its impacts on individuals residing within long term care facilities (LTCFs) such as rehabilitation centers and nursing homes. LTCF residents tend to possess several risk factors for more severe SARS-CoV-2 outcomes, including advanced age and multiple comorbidities. Indeed, residents of LTCFs represent approximately 40% of SARS-CoV- 2 deaths in the United States.

**Methods:** To assess the prevalence and incidence of SARS-CoV-2 among LTCF workers, determine the extent of asymptomatic SARS-CoV-2 infection, and provide information on the genomic epidemiology of the virus within these unique care settings, we collected nasopharyngeal swabs from workers for 8-11 weeks at six Colorado LTCFs, determined the presence and level of viral RNA and infectious virus within these samples, and sequenced 54 nearly complete genomes.

**Findings:** Our data reveal a strikingly high degree of asymptomatic/mildly symptomatic infection, a strong correlation between viral RNA and infectious virus, prolonged infections and persistent RNA in a subset of individuals, and declining incidence over time.

**Interpretation:** Our data suggest that asymptomatic SARS-CoV-2 infected individuals contribute to virus persistence and transmission within the workplace, due to high levels of virus. Genetic epidemiology revealed that SARS-CoV-2 likely spreads between staff within an LTCF.

**Funding:** Colorado State University Colleges of Health and Human Sciences, Veterinary Medicine and Biomedical Sciences, Natural Sciences, and Walter Scott, Jr. College of Engineering, the Columbine Health Systems Center for Healthy Aging, and the National Institute of Allergy and Infectious Diseases.

**Research in Context:** *Evidence before this study:* We searched PubMed and Google Scholar on April 15, 2020 for manuscripts published in 2020 with the key words “SARS-CoV-2 OR COVID-19 AND Long-Term Care Facility AND Surveillance OR Screening. We did not restrict our search to the English language. Our search retrieved two reports of original research. The relevant publications described transmission and course of infection among residents in LTCFs. Of particular relevance was that large quantities of SARS-CoV-2 viral RNA could be detected in asymptomatic, presymptomatic and symptomatic residents, providing early evidence of the heterogeneity of infection characteristics among residents at LTCFs. A significant number of LTCF residents were presymptomatic with symptoms emerging 7 days after initial detection of viral RNA, indicating a longer than expected latency period. Therefore, symptomatic screening for early detection and resultant mitigation response was likely to be ineffective in preventing transmission among residents of LTCFs. There were no reports involving longitudinal surveillance testing of LTCF staff.

*Added value of this study:* While prior studies reported results of facility-wide (residents and staff) testing for SARS-CoV-2 and describe transmission dynamics among residents of LTCFs, no prior data was available describing the longitudinal characteristics of SARS-CoV-2 dynamics among staff working at LTCFs during a time period where “shelter-in-place” public guidance was in effect. During this time period, LTCF residents were largely isolated, however staff (those with both direct care and those without direct contact) were permitted to leave and return to work daily. We were therefore interested in this broad staff cohort specifically because they represent a significant and ongoing potential source of transmission within LTCFs. RT-qPCR testing for SARS-CoV-2 was performed weekly on 544 staff in six LTCFs over an 8-11-week period. Symptom data were collected and site-specific prevalence at study onset and incidence rate over time were calculated to explore the influence of identifying and removing asymptomatic SARS-CoV-2-infected individuals from the workplace.

*Implications of all the available evidence:* Our results document a surprising degree of asymptomatic/mildly symptomatic infection among apparently healthy staff, and extreme variation in SARS-CoV-2 prevalence and incidence among staff between different facilities. Plaque assay revealed a strong relationship between vRNA and infectious virus in nasopharyngeal swab material, indicating the asymptomatic or mildly symptomatic individuals are infectious. Moreover, phylogenetic analysis of SARS-CoV-2 sequences collected from LTCF staff suggest that the predominant transmission pattern is between staff members within facilities, and that individual unrelated community import events are less common. Finally, decreasing prevalence over time within facilities where longitudinal surveillance testing was performed suggests that identifying and isolating positive staff may serve as part of an effective mitigation program to prevent or curtail transmission among staff within LTCFs.

## Introduction

The highly infectious SARS-CoV-2 virus threatens the stability of healthcare systems around the world. Long term care facilities (LTCFs), due to their communal nature, the limited mobility of their inhabitants and the propensity of residents to have underlying health conditions, have become significant venues of virus transmission [1]. The COVID-19 pandemic has resulted in disproportionally high morbidity and mortality among residents in LTCFs. As of October 10, 2020, the Centers for Medicare and Medicaid Services reported over 84,000 deaths due to COVID-19 in U.S. LTCFs, representing over 38% of COVID-19-related deaths [2, 3]. In the U.S., the first recorded SARS-CoV-2 outbreak occurred in a LTCF in Washington as early as February [4]. Since then, every state has recorded outbreaks in LTCFs, and in 14 states LTCF deaths account for over 50% of all COVID-19 deaths [3]. The high mortality associated with SARS-CoV-2 infection within LTCFs is principally due to the risk profiles of residents residing in communal care settings, including advanced age and pre-existing comorbidities, such as heart disease and diabetes mellitus [5-7].

Accordingly, strategies to mitigate SARS-CoV-2 transmission to LTCF residents have included restricting visitation, cessation of group activities and dining, and confinement to individual living quarters [8-11]. While LTCF residents have been largely isolated from external visitation,staff are permitted contact provided they have passed a daily screening process to asses for fever, COVID-19 respiratory symptoms or known exposure [12].These staff have the potential to import the virus into facilities, resulting in spread to residents, other workers, and back to the outside community [1]. While symptom screening can reduce virus spread, a significant fraction of individuals infected with SARS-CoV-2 have a lengthy latency period prior to exhibiting COVID-19 symptoms, and many remain asymptomatic throughout the course of infection [13- 18]. Therefore, pre-symptomatic, asymptomatic and mildly symptomatic LTCF staff are a potential source of transmission within LTCFs and are thus an attractive focus for interventions directed at suppressing infections within these facilities [15, 16, 19-23].

While there are a growing number of studies measuring SARS-CoV-2 infection within LTCF residents, there are limited studies focusing on longitudinal surveillance of LTCF asymptomatic staff [24]. In Colorado, cases linked to LTCFs account for over 49% of all COVID-19 deaths [2, 3]. To evaluate the impact of staff on virus introduction into LTCFs, we tested staff at six Colorado LTCFs for SARS-CoV-2. Staff were enrolled and sampled by nasopharyngeal swab weekly for 8-11 consecutive weeks. Samples were assayed for virus by RT-qPCR and plaque assay, and individuals with evidence of infection were instructed to self-quarantine for ten days. Return to work required absence of fever for the final three days of isolation. Using data on staff infection, site-specific prevalence at study onset and incidence rate over time were calculated. Viral genomes were sequenced to assess viral genetic diversity within and between LTCFs.

Our results document a surprising degree of asymptomatic/mildly symptomatic infection among apparently healthy staff, and extreme variation in SARS-CoV-2 prevalence and incidence between different facilities, similar to what has been observed at other LTCFs [15, 16, 19, 22]. We documented a range of infection courses, including acute (1 week), prolonged (4+ weeks), and recrudescent. Sequencing studies lend support to the observation that transmission may occur within LTCFs and, combined with the epidemiologic and other data provided here, highlight the importance of testing and removing virus-positive workers in order to protect vulnerable LTCF residents. Data obtained from longitudinal surveillance studies provide crucial information about infectious disease transmission dynamics within complex workforces and inform best practices for preventing or mitigating COVID-19 outbreaks within LTCFs.

## Materials and Methods

### Study sites

Staff at LTCFs provided consent to participate in this study. Nasopharyngeal (NP) swabs, or saliva (only sampled once at two facilities when swabs were unavailable) were collected weekly for 8-11 weeks. Participants provided date of birth and job code but were otherwise de-identified. This study was reviewed and approved by the Colorado State University IRB under protocol number 20-10057H. Participants were promptly informed of test results and when positive, instructed to self-isolate for ten days. Return to work required absence of fever or other symptoms for the final three days of isolation.

### Sample collection

Nasopharyngeal swabs were collected by trained personnel. Swabs were placed in a conical tube containing 3ml viral transport media (Hanks Balanced Salt Solution, 2% FBS, 50mg/ml gentamicin, 250ug/ml amphotericin B/fungizone). Saliva was collected by repeatedly spitting through a straw into a sterile tube.

### RNA extraction

Tubes containing NP swabs were vortexed and centrifuged to pellet debris. RNA was extracted from supernatant with the Omega Mag-Bind Viral DNA/RNA 96 Kit using 200ul of input sample on a KingFisher Flex magnetic particle processor according to the manufacturers’ instructions.

### qRT-PCR

One-step reverse transcription and PCR was performed using the EXPRESS One-Step SuperScript qRT-PCR Kit (ThermoFisher Scientific) per the manufacturers’ instructions. N1, N2, and E primer/probes were obtained from IDT and described elsewhere [25-27]. RNA standards for nucleocapsid (N) and envelope (E) were provided by Dr. Nathan Grubaugh of Yale University and used to determine copy number [26]. Samples were screened with N1 primer/probes, and those with a cycle threshold (CT) less than 38 were tested for N2 and E vRNA.

### Plaque assay

Plaque assays were performed on African Green Monkey Kidney (Vero) cells (ATCC CCL-81) according to standard methods [28]. Briefly, 250uL of serially diluted samples were inoculated onto cell monolayer for one hour. After incubation, cells were overlaid with tragacanth medium, incubated for two days, fixed and stained with 30% ethanol and 0.1% crystal violet. Plaques were counted manually.

### Incidence estimation

The rate at which staff acquired infections was estimated as the number of new infections per 100 workers per week at each facility from week 2 through the end of the study. Staff were classified as having an incident infection if they tested positive for the first time following a negative test one- or two-weeks prior and if they had not previously tested positive for SARS-CoV-2 in our study. The population at risk included all staff who had not yet been infected, to our knowledge, and who tested negative in week one of the study.

### Symptom reporting

Symptom data were collected and managed with REDCap electronic data capture tools hosted at the Colorado Clinical and Translational Sciences Institute (CCTSI) at University of Colorado Anschutz Medical Campus [29, 30]. Survey administrators accessed the survey on a portable tablet computer, entered a participant-specific case number, and provided a verbal introduction. Participants were asked to enter responses to questions concerning symptoms, symptom severity, comorbidities, household size, general characteristics (height, weight, etc.), smoking habits, inhaled medication use, and potential exposure to SARS-CoV-2. Symptom severity and exposure questions were phrased to encompass a range of time from mid-March to late-June. Survey participants were asked to recall symptoms coinciding with this time period.

### Next-generation sequencing and analysis

cDNA was generated using SuperScript IV Reverse Transcriptase enzyme (Invitrogen) with random hexamers. PCR amplification was performed using ARTIC network V2 or V3 tiled amplicon primers in two separate reactions by Q5 High-Fidelity polymerase (NEB) as previously described [31]. First-round PCR products were purified using Ampure XP beads (Beckman Coulter). Libraries were prepared using the Nextera XT Library Preparation Kit (Illumina) according to manufacturer protocol. Unique Nextera XT i7 and i5 indexes for each sample were incorporated for dual indexed libraries. Indexed libraries were again purified using Ampure XP beads. Final libraries were pooled and analyzed for size distribution using the Agilent High Sensitivity D1000 Screen Tape on the Agilent Tapestation 2200. Final quantification was performed using the NEBNext Library Quant Kit for Illumina (NEB) according to manufacturer protocol. Libraries were sequenced on the Illumina MiSeq V2 using 2 x 250 paired-end reads.

Sequencing data were processed to generate consensus sequences for each viral sample. MiSeq reads were demultiplexed, quality checked by FASTQC, paired-end reads were processed to remove Illumina primers and quality trimmed with Cutadapt; duplicate reads were removed. Remaining reads were aligned to SARS-CoV-2 WA1-F6/2020 reference sequence by Bowtie2 (GenBank: MT020881.1). Alignments were further processed, quality checked using Geneious software, consensus sequences were determined, and any gaps in sequences were filled in with the reference sequence or cohort specific consensus sequence. Consensus sequences were aligned in Geneious and a maximum-likelihood tree generated using PhyML in Geneious with the Wuhan-Hu-1 reference sequence (GeneBank: MN908947.3) as an outgroup and 100 bootstrap replicates.

## Results

### Cohort characteristics

From March 26 to June 23, 2020, we tested 544 staff from six LTCFs (**Table 1**). Of these participants, 91 (16.7%) tested positive for SARS-CoV-2 viral RNA (vRNA) at least once during the study. We tested 3, 754 samples total, of which 179 were positive for vRNA (4.77% of total samples).

**Table 1.**
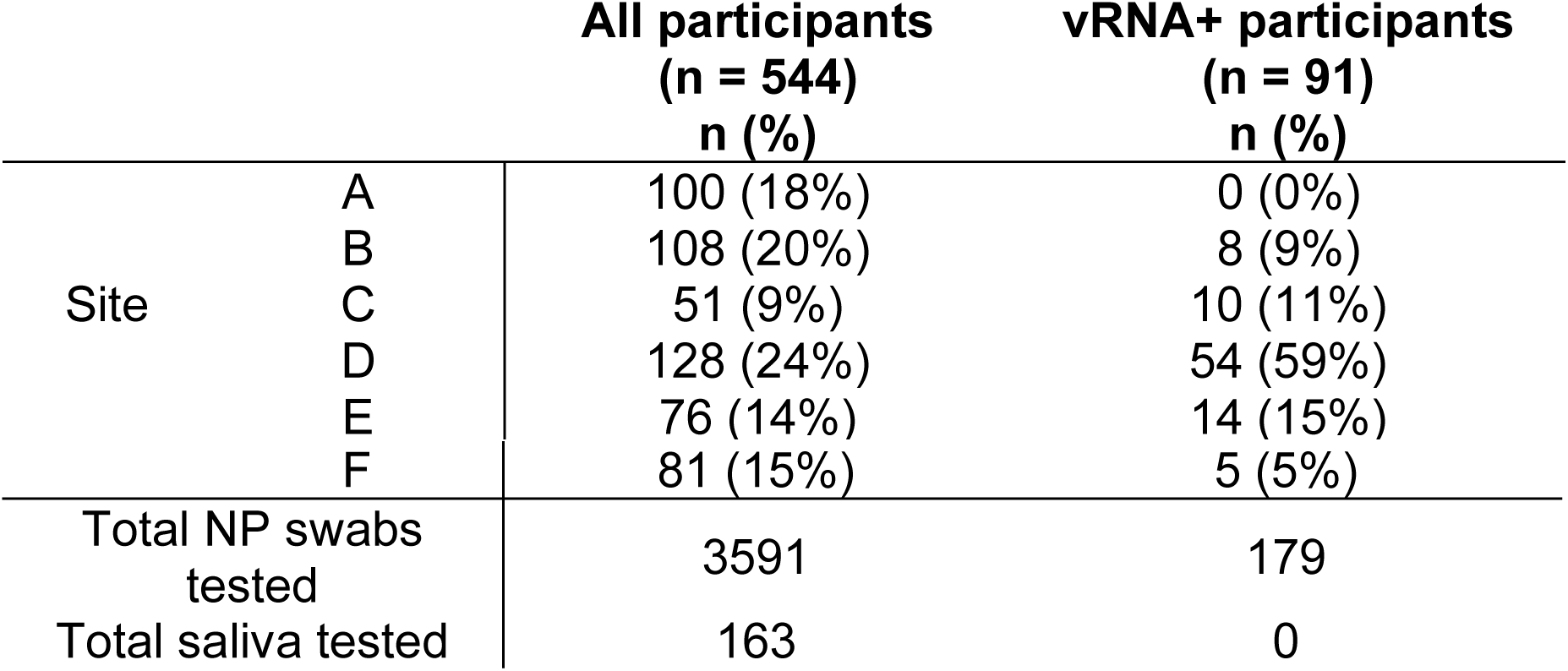
Colorado LTCF cohort characteristics.

### Viral load, prevalence and incidence rate vary across LTCFs

Viral RNA levels and the prevalence of vRNA-positive swabs varied each week by site (**Fig. 1A & B**). Staff at Site A remained uninfected throughout the entire 8-week study period, whereas 31% of individuals at site D were infected on week two. All sites showed a decline in SARS-CoV-2 prevalence over the course of the study (**Fig. 1B**). SARS-CoV-2 incidence also varied across sites (**Fig. 1C**). At site D, which had the highest SARS-CoV-2 prevalence, the initial incidence was also high (13.6 cases per 100 person-weeks) but declined over time. At sites C and F, the incidence reached zero by week 3, however both sites had a small number of incident cases in later weeks. Sites B and E, which had low prevalence in week 1, saw an increase in cases. At site B, incident infections were detected after three weeks. Infections were observed in all job classes, including those with typically high patient contact (e.g. nursing) and low patient contact (e.g. maintenance) (**Table 2**). The highest odds ratios for infection occurred in housekeeping, nursing and staff in other jobs, while the lowest were in administration, therapy and dietary staff (**Table 2**).

**Table 2.** Analysis of infections in LTCF staff by job code. The distribution of infections by job code among 435 staff at LTCFs where SARS-CoV-2 was detected during the study period.

| Job code | Number tested | % positive* | Unadjusted OR (95% CI) | Adjusted OR (95% CI) |
| --- | --- | --- | --- | --- |
| Administration | 53 | 11.3 | 1.00 (ref) | 1.00 (ref) |
| Nursing | 180 | 24.4 | 2.53 (1.01, 6.33) | 2.79 (1.07, 7.32) |
| Housekeeping | 96 | 14.6 | 1.34 (0.48, 3.71) | 4.69 (1.39, 15.84) |
| Dietary | 36 | 19.4 | 1.89 (0.58, 6.18) | 1.55 (0.45, 5.34) |
| Therapy | 24 | 4.2 | 0.34 (0.04, 3.00) | 0.47 (0.05, 4.45) |
| Other** | 46 | 34.8 | 4.18 (1.47, 11.87) | 4.91 (1.61, 14.97) |

\*Analysis looks at the percent of workers that tested positive at least once during the study period. Analysis is limited to the five sites where SARS-CoV-2 was detected (B, C, D, E, F). Unadjusted odds ratios were estimated using logistic regression, adjusted analyses included a dummy variable for site. \*\*Other jobs include physician/provider, maintenance, social services, transport, and activities.

**Figure 1.**
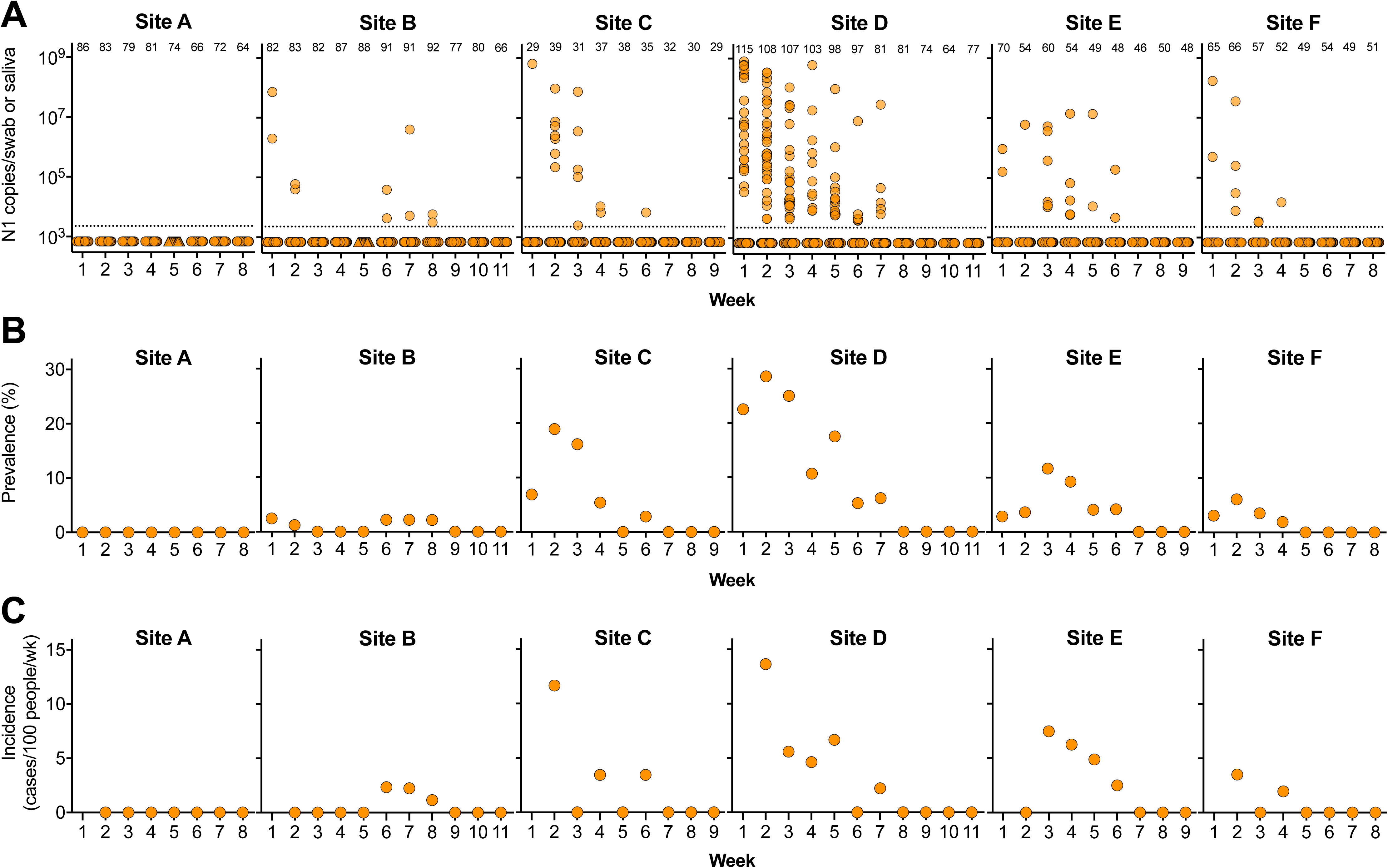
SARS-CoV-2 infection in six Colorado LTCFs. **A)** SARS-CoV-2 N1 vRNA levels in nasopharyngeal swabs (circle) or saliva (triangle). Y-axis represents N1 copies/swab or saliva. Dotted line indicates limit of detection. Numbers across the top indicate number of samples tested each week. **B)** Prevalence of SARS-CoV-2 each week at each site (percent of samples with detectable N1 vRNA out of total number tested). **C)** Incident cases were defined as individuals who tested positive for N1 vRNA for the first time and had tested negative for infection one or two weeks prior. Not shown are prevalent infections among workers tested for the first time in week two.

### Relationship between viral RNAs and infectious virus in nasopharyngeal swabs

Swabs with SARS-CoV-2 N1 vRNA were tested for N2- and E-containing viral transcripts (**Fig. 2A**). We observed high concordance between levels of N1 and N2 vRNA, with a median genome to genome ratio of 1.2 (**Fig. 2B**). E vRNA levels were lower and less detectable than either N1 or N2 (**Fig. 2A**), consistent with coronavirus replication, resulting in higher genome ratios (**Fig.2B**). Samples with detectable N1 vRNA were also tested for infectious virus. We found a strong positive relationship between vRNA and infectious virus in swab material (**Fig. 2C**). Infectious virus was rarely detected in individuals with fewer than 10^5^ N1 vRNA copies. However, there were some samples with high levels of vRNA (∼10^7^ copies) with undetectable infectious virus. Virus specific infectivity varied depending on the region of the genome analyzed (**Fig. 2D**).

**Figure 2.**
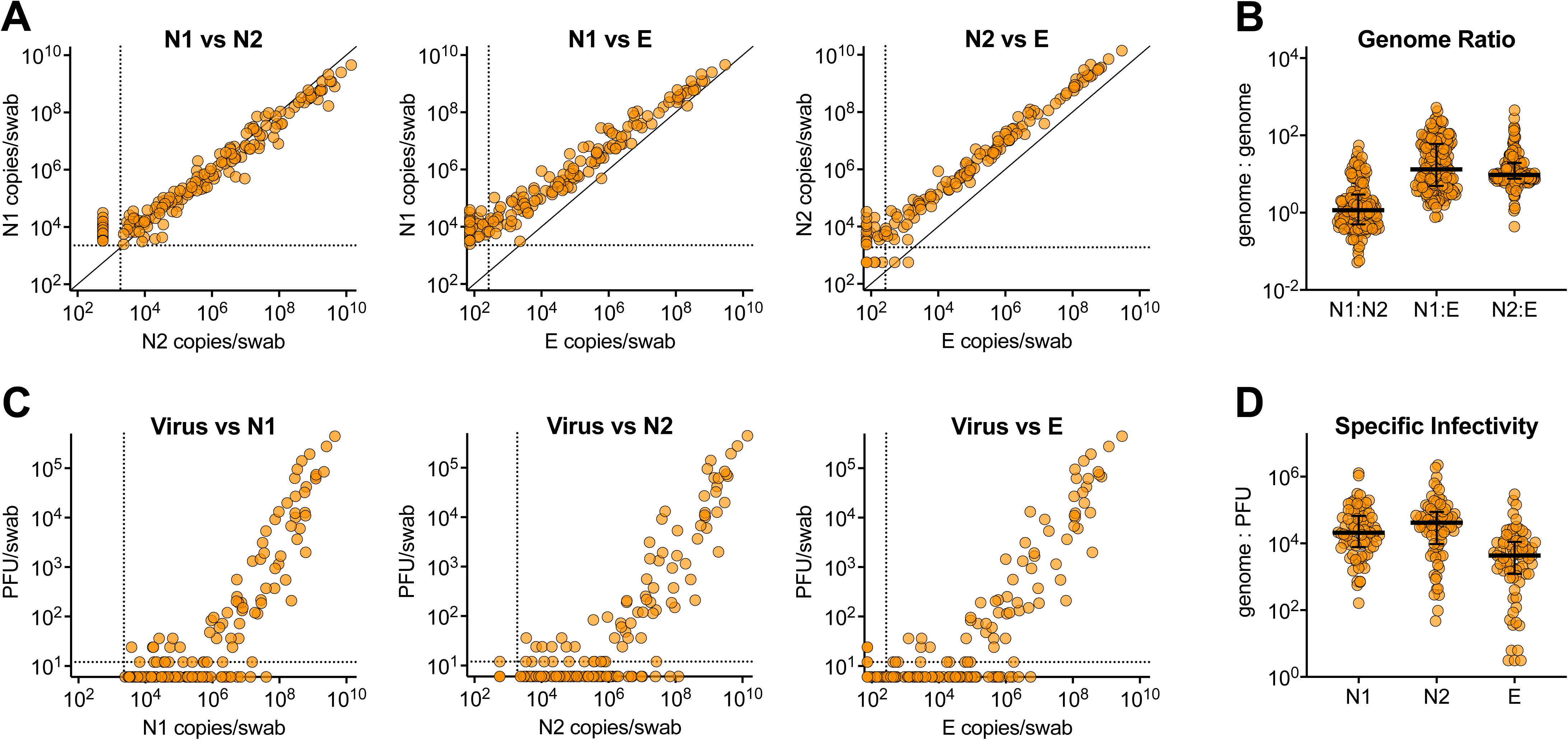
Relationship between SARS-CoV-2 viral RNA and infectious virus. Samples with detectable SARS-CoV-2 N1 vRNA were evaluated for N2 and E vRNA and infectious virus. **A)** Relationship between levels of N1, N2 and E vRNA transcripts. **B)** Genome:genome ratios between N1:N2, N1:E and N2:E (median with interquartile range). **C)** Relationship between levels of infectious virus and N1, N2, and E vRNA levels. **D)** Specific infectivity (genome:PFU ratio) of infectious virus relative to N1, N2 and E transcripts (median with interquartile range). Dashed lines represent limits of detection. PFU, plaque forming units.

### SARS-CoV-2 infection and vRNA levels are not related to age, BMI, sex or job code

Age, body mass index (BMI), sex and smoking habits have been implicated in SARS-CoV-2 infection and disease outcomes [32-38]. We detected no significant differences between these variables among vRNA-negative and vRNA-positive individuals (**Table 3**). Viral RNA level from N1- positive samples was not dependent on age, BMI, sex, smoking habits or job code (**SFig. 1**).

**Table 3.** Age, BMI and smoking status among cohort subset.

|  | vRNA- | vRNA+ | p-value |
| --- | --- | --- | --- |
| Age, mean (range) | 41 (17-76)<br>(n = 454) | 41 (16-72)<br>(n = 91) | 0.7645† |
| BMI, mean (range) | 28.7 (17.8-46.6)<br>(n = 190) | 28.2 (20.8-43.0)<br>(n = 51) | 0.3265† |
| Current smokers | 21.2% (40/190) | 16.3% (8/49) | 0.5516‡ |
| Former smokers* | 20.0% (28/190) | 24.5% (12/49) | 0.1315‡ |
| Marijuana smokers | 5.3% (10/188) | 6.1% (3/49) | 0.7348‡ |
| Tobacco-based vape product users | 6.3% (12/189) | 4.2% (2/48) | 0.7412‡ |

\*Former smoker refers to those who answered 'Yes' to 'are you a former smoker' and 'No' to 'Do you currently smoke cigarettes'.
†T-test, ‡Fisher's Exact Test

### Symptom status differs based on SARS-CoV-2 infection status

A subset of study participants (n = 191 vRNA-, n = 51 vRNA+), responded to a survey to capture recollection of eleven COVID-19-related symptoms during the study period [39] (**Table 4**). All symptoms were significantly more frequent among infected participants. Cough and fever >100.4°F, two symptoms commonly used for COVID screening, were reported in 48% and 24% of infected participants, as compared to 14.3% and 7.4% in uninfected individuals. Other symptoms such as the loss of taste and smell (ageusia and anosmia), were significantly associated with SARS-CoV-2 infection (reported in 2.1% of vRNA-negative and 51.0% of vRNA-positive individuals).

**Table 4.**
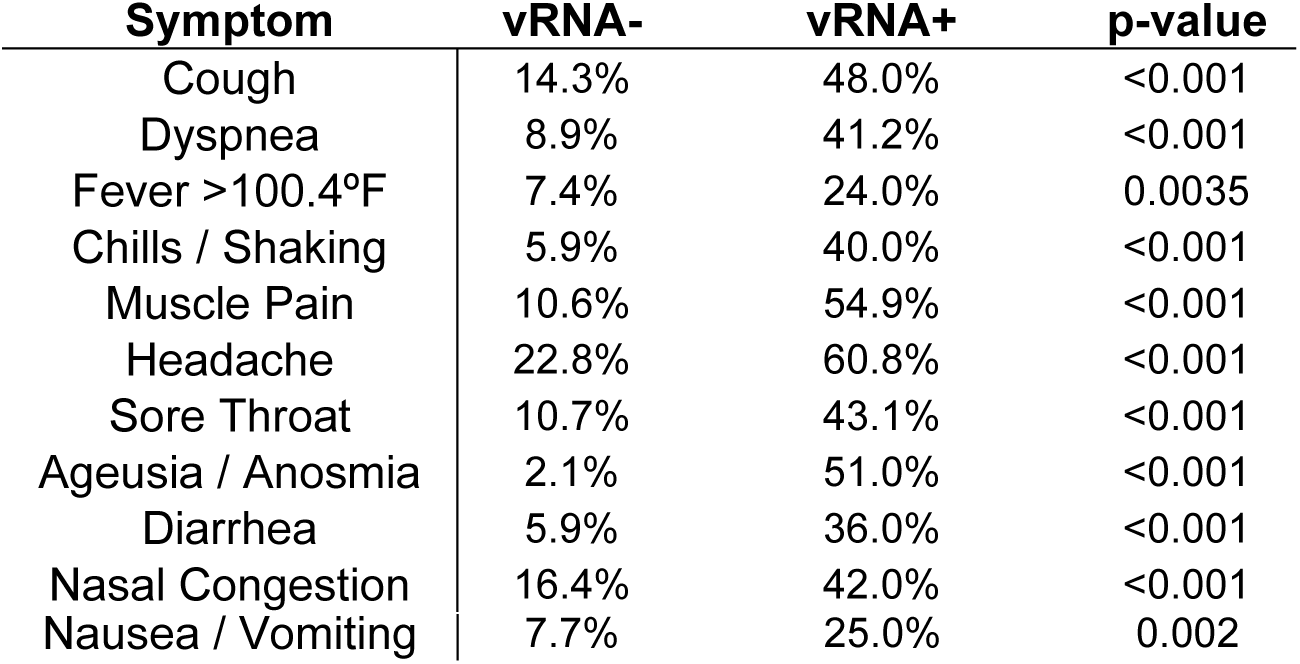
Symptom status among vRNA-negative and positive individuals.

### Symptom status and severity is related to SARS-CoV-2 infection

vRNA-positive individuals recalled more symptoms than vRNA-negative individuals (p<0.001) (**Fig. 3A**). Almost 80% of vRNA-negative individuals experience 0-1 symptoms, whereas vRNA-positive individuals evenly recalled a range of symptoms (**Fig. 3B**). 27% of vRNA-positive individuals reported zero symptoms, and 41% reported 2 or fewer symptoms (**Fig. 3C**). Severity was scored (0-no symptom, 1-mild, 2-moderate, 3-severe) for each symptom, and symptom score was compared between vRNA-negative and positive individuals. Average symptom score was significantly higher in vRNA-positive individuals (p<0.001) (**Fig. 3D**). Over 70% of vRNA-negative individuals had a symptom severity score of 1 or less, whereas vRNA-positive individuals had an evenly broad range of scores (**Fig. 3E**). Within vRNA-positive individuals, total symptom score was not correlated with N1 vRNA levels (**Fig. 3F**). N1 vRNA levels were stratified by severity for each symptom. N1 vRNA did not predict the severity of any symptom independently (**S2Fig)**.

**Figure 3.**
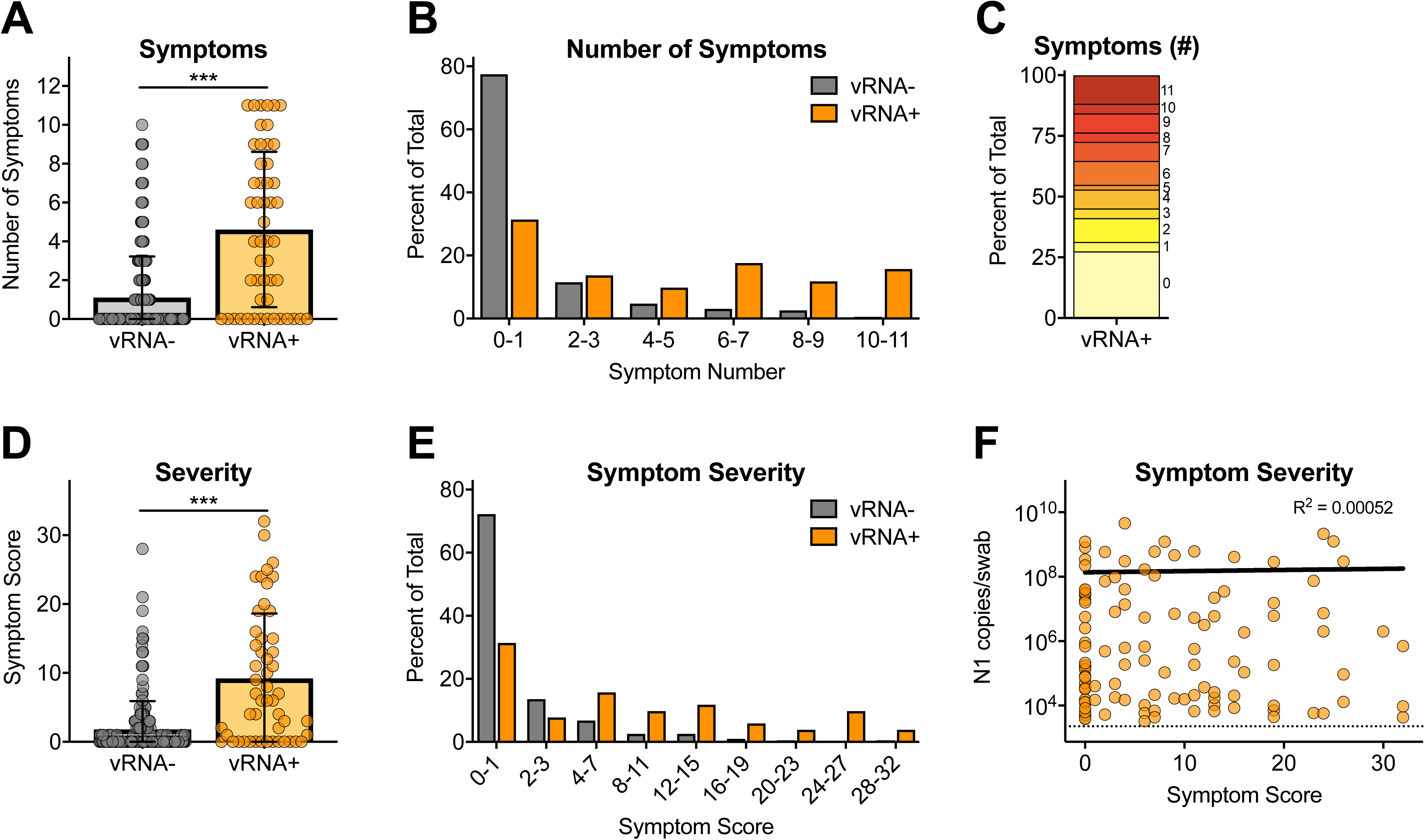
SARS-CoV-2 symptom status, severity and relationship to viral RNA. **A)** Number of symptoms reported by vRNA- and vRNA+ participants (mean ± SD). **B)** Percentage of vRNA- and vRNA+ individuals stratified by number of symptoms. **C)** Percentage of vRNA+ survey participants reporting total number of symptoms. **D)** Cumulative symptom score (not reported = 0, mild = 1, medium = 2, severe = 3) for all 11 symptoms stratified by vRNA- and vRNA+ participants (mean ± SD). **E)** Percentage of vRNA- and vRNA+ individuals stratified by symptom score. **F**) Relationship between cumulative symptom score and N1 vRNA levels (semilog nonlinear regression line fit). *** p<0.0001 Mann-Whitney unpaired non-parametric test.

### Participants experienced acute, prolonged and resurgent SARS-CoV-2 infections

Within the cohort and study period, we observed a range of infection courses (**Fig. 4A-E**). Individuals who were positive for a single week included those with low levels of vRNA and no detectable infectious virus (B150), to those with high levels of both vRNA and infectious virus (F058) (**Fig. 4A**). Individuals who were positive for multiple consecutive weeks often had high levels of virus on their first positive test which decreased in subsequent weeks (**Fig. 4B-D**). There were also individuals with positive SARS-CoV-2 tests followed by 1-3 weeks of negative tests, before vRNA was again detected (**Fig. 4E**). Individuals with incident infections during the course of the study, with negative tests before and after positives, were stratified based on the number of consecutive vRNA-positive weeks (**Fig. 4F**). Those who were vRNA-positive for a single week tended to have low N1 levels and rarely had infectious virus (**Fig. 4F**). Virus levels in infections that lasted 2-4 weeks, were generally highest on the first week and subsequently decreased (**Fig. 4F**). Individuals with post-negative positive tests (positive after 1-3 weeks of negative tests following initial infection), were associated with very low levels of vRNA and rarely infectious virus (**Fig. 4F**).

**Figure 4.**
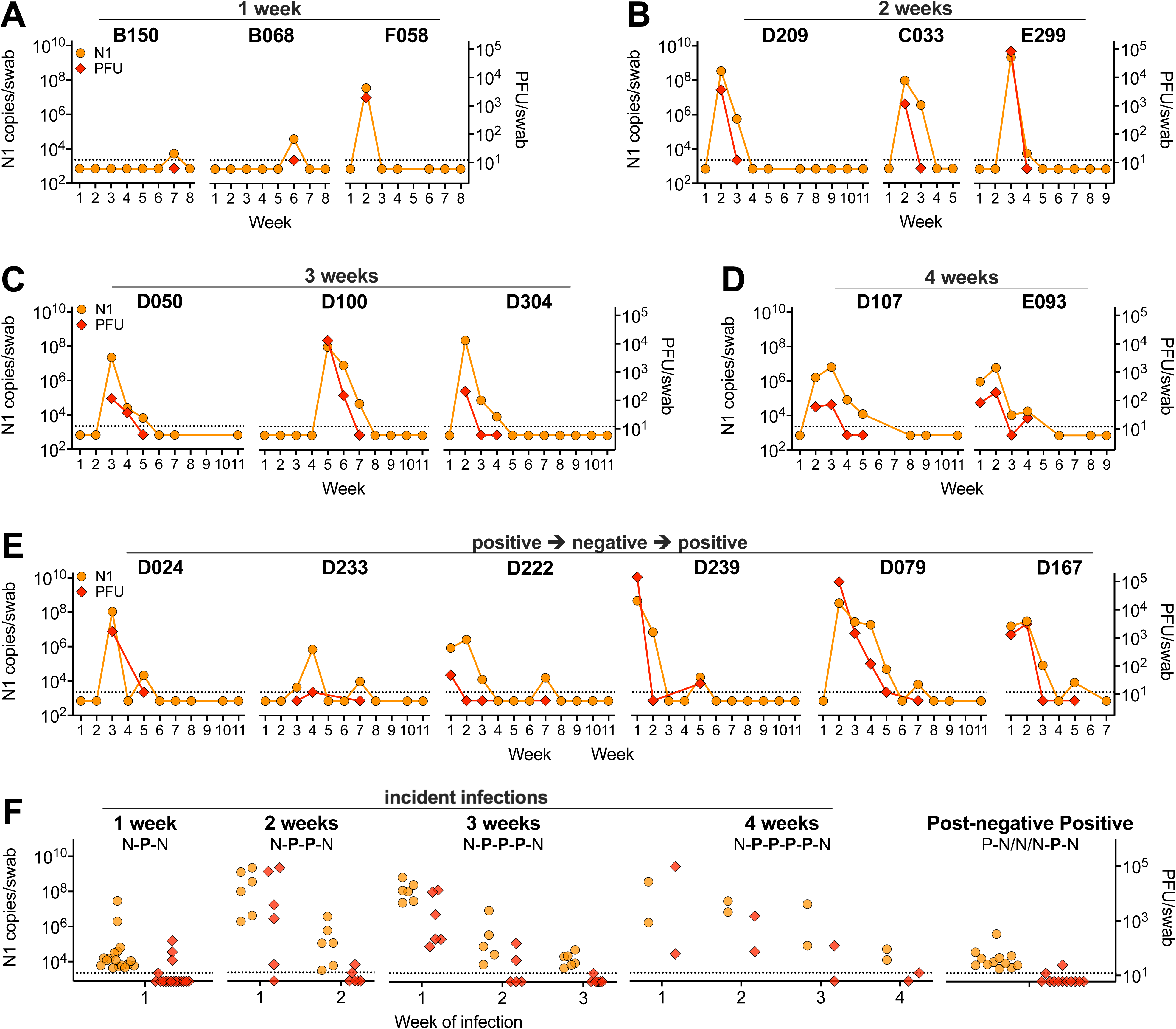
Individual infection courses and virus levels. Viral N1 RNA (left axis) and infectious virus (right axis) in select individuals with detectable N1 for **A)** one, **B)** two, **C)** three, or **D)** four consecutive weeks. **E)** Examples of individuals with detection of N1 vRNA after a period of undetectable N1 following initial infection. **F)** N1 vRNA and infectious virus by week of infection is plotted for individuals with incident infectious during the course of the study, with negative (N) tests immediately before and after positive (P) tests, stratified by the length of infection (one, two, three or four consecutive positive weeks) and those who experienced a post-negative positive test (following 1-3 negative weeks). Dashed line represents limit of detection, samples not detected plotted at half the limit of detection. PFU, plaque forming units.

### Phylogenetic analysis of SARS-CoV-2 sequences from LTCFs

54 partial genome sequences were obtained from individuals with infections during the study (**Fig. 5**). Mean genome coverage was 29,317nt (range = 24,076-29,835) and mean coverage depth was 640 reads per position (range = 344-2,138). Gaps in sequencing alignment due to ARTIC V2/V3 primer incompatibilities were filled in with the reference strain MT020881.1. The LTCF sequences were aligned to a reference strain from early in the U.S. outbreak (WA1-F6), four Colorado strains (CO-CDC), and strains from California (USA-CA1), New York (USA/NY) and Wuhan (Wuhan-Hu-1). The tree was reasonably resolved into multiple clusters with moderate bootstrap support (i.e. >50%). The largest cluster is composed exclusively of sequences obtained from individuals at site D (**Fig. 5**, lower part of tree). Sequences from sites C (red) and E (orange) primarily cluster amongst themselves, however there are site C sequences within the D clusters as well. The single sequence from site B (B137_05/08/20), is most similar to site C sequences.

**Figure 5.**
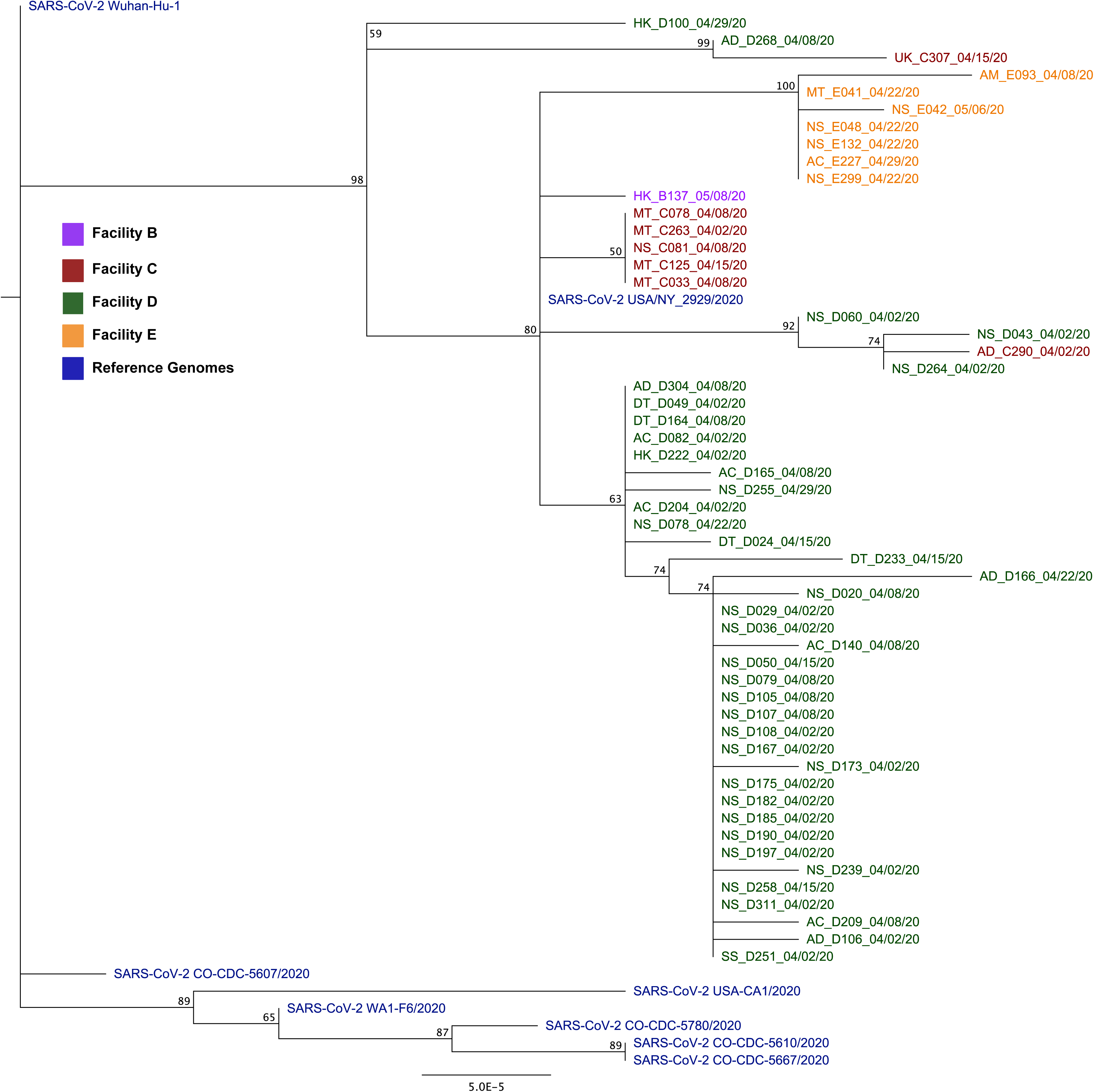
Phylogenetic analysis of SARS-CoV-2 genomes collected from Colorado LTCFs. **A)** PhyML tree constructed using Tamura-Nei distance model including both transitions and transversions in Geneious Prime. Node numbers indicate bootstrap confidence based on 1000 replicates. Distance matrix was computed, and the tree was visualized in Geneious Prime. Letters at the beginning of taxon names represent job code (AC-activities, AD-administrative, AM-admissions, DT-dietary, MT-maintenance, NS-nursing, SS-social services, UK-unknown), and A-E letter indicate site of origin. Numbers after underscore indicate the date of sample collection. Reference sequences and four Colorado-derived sequences were obtained from NCBI. **B)** Map of the LTCFs’ relative geographic locations and distances from one another.

## Discussion

LTCFs are increasingly recognized as high-risk for SARS-CoV-2 transmission [12, 19, 23]. Because of their disproportionate contribution to the burden of COVID-19 mortality [2, 3], they also represent an attractive target for surveillance testing [11]. Consistent with other LTCF cohorts [15, 16, 20], our data clearly demonstrate the potential for large numbers of staff at LTCFs to be asymptomatically/presymptomatically infected and for the concentration of infection to vary widely across facilities. One facility had no positive staff, while others had up to 30% of staff test positive within the same sampling period. The steady decline in new infections in facilities with the highest initial infection prevalence following removal of SARS-CoV-2-positive staff from the workplace is encouraging and hints at the potential impact of longitudinal surveillance. The detection of incident infections at facility B, after three weeks of negative tests underscores the on-going threat of infections in worker populations. These results clearly demonstrate that infected staff may be common in specific LTCFs [15-17, 19].

Because coronavirus genome replication creates an abundance of sub-genomic N-containing transcripts [40], it is therefore not surprising that higher levels of N transcripts are detected compared to E vRNA. We found that viral RNA was strongly correlated with infectious virus (samples with high levels of vRNA tended to have high levels of infectious virus, whereas lower vRNA levels often had undetectable levels of infectious virus). Importantly, this demonstrates that individuals with high levels of vRNA are likely infectious to others [41-43]. We also detected infectious virus in asymptomatic individuals, and at time points later than other reports, suggesting that presence and duration of infectious virus varies greatly by individual [44]. Our data supports the observation that seemingly healthy staff can harbor high levels of infectious virus in the absence of clinical disease and may therefore contribute to transmission of SARS-CoV-2.

The impact of age, sex, BMI, race, ethnicity, and other patient characteristics on SARS-CoV-2 infection and disease outcomes are not well defined [32-37]. Within our cohort, we detected no relationship between any of these factors and RNA load, symptom number or severity.

Additionally, while symptom status and severity are strongly correlated to positive SARS-CoV-2 results, viral load is not correlated with either status or severity. Notably, others have found that symptomatic hospitalized patients have lower virus levels than non-hospitalized peers [45]. Together, these results suggest that other host or viral factors likely impact virus level and clinical presentation.

The longitudinal design of this study permitted characterization of individuals’ full infection courses, including those who were positive for 1-5 consecutive weeks. In most cases, viral load was highest in the first week, then declined. Consistent with other reports [46-49], we observed individuals with positive tests after apparent clearance of the initial infection. While it is possible that these individuals were re-infected immediately after clearing their initial infection, we find that unlikely [50, 51]. Instead, this may be due to host factors that lead to temporary suppression of virus within the nasopharynx, or an improper swab collection that failed to capture sufficient material for detection [52]. Importantly, the post-negative positive samples contained low levels of vRNA, and low or undetectable infectious virus. These data highlight the heterogeneity of human SARS-CoV-2 infection, and the need to further understand host and viral factors that govern infection and clearance.

Virus sequencing provides insights into SARS-CoV-2 transmission [24]. Our data encompasses 54 genomes obtained from four sites. Strikingly, the viruses primarily cluster by facility, suggesting local transmission among staff at each site. It is possible there are also community-acquired infections which are introduced to the facilities, which could explain highly similar virus sequences at multiple sites. Data on the degree of viral genetic diversity in the larger community would add significant power to our ability to discriminate between these two non-mutually exclusive scenarios. Additional comparisons to existing SARS-CoV-2 sequences would also help elucidate introduction and spread within the facilities and Colorado as a whole [31].

Overall, our study highlights the high SARS-CoV-2 infection rates within staff at LTCFs. Identifying and isolating these infected and infectious individuals, may serve as an effective mitigation strategy. While our work focused on LTCFs, this approach could be applied to other communal living settings (correctional facilities, factories, etc.).

## Data Availability

N/A. Sequence data will be deposited and made publicly available once it is completed. Files in their current state are available from the authors upon request.

## Acknowledgements

This work was supported by funds donated by the Colorado State University Colleges of Health and Human Sciences, Veterinary Medicine and Biomedical Sciences, Natural Sciences, and Walter Scott, Jr. College of Engineering, and the Colorado State University Columbine Health Systems Center for Healthy Aging. KMQ was supported by a fellowship from the National Institute of Allergy and Infectious Diseases, National Institutes of Health under grant number F32AI150123-01. The authors also gratefully acknowledge the CSU Veterinary Diagnostic Laboratory for diagnostic support, Carolina Mehaffy for courier support, and the participation of the workers in the facilities that participated in this study, without which it could not have been completed. The funding sources had no role in the writing of this manuscript of the decision to submit it for publication. None of the authors have been paid to write this publication. The authors declare no conflicts of interest.

## Supplemental Legends

**Supplemental Figure 1.**
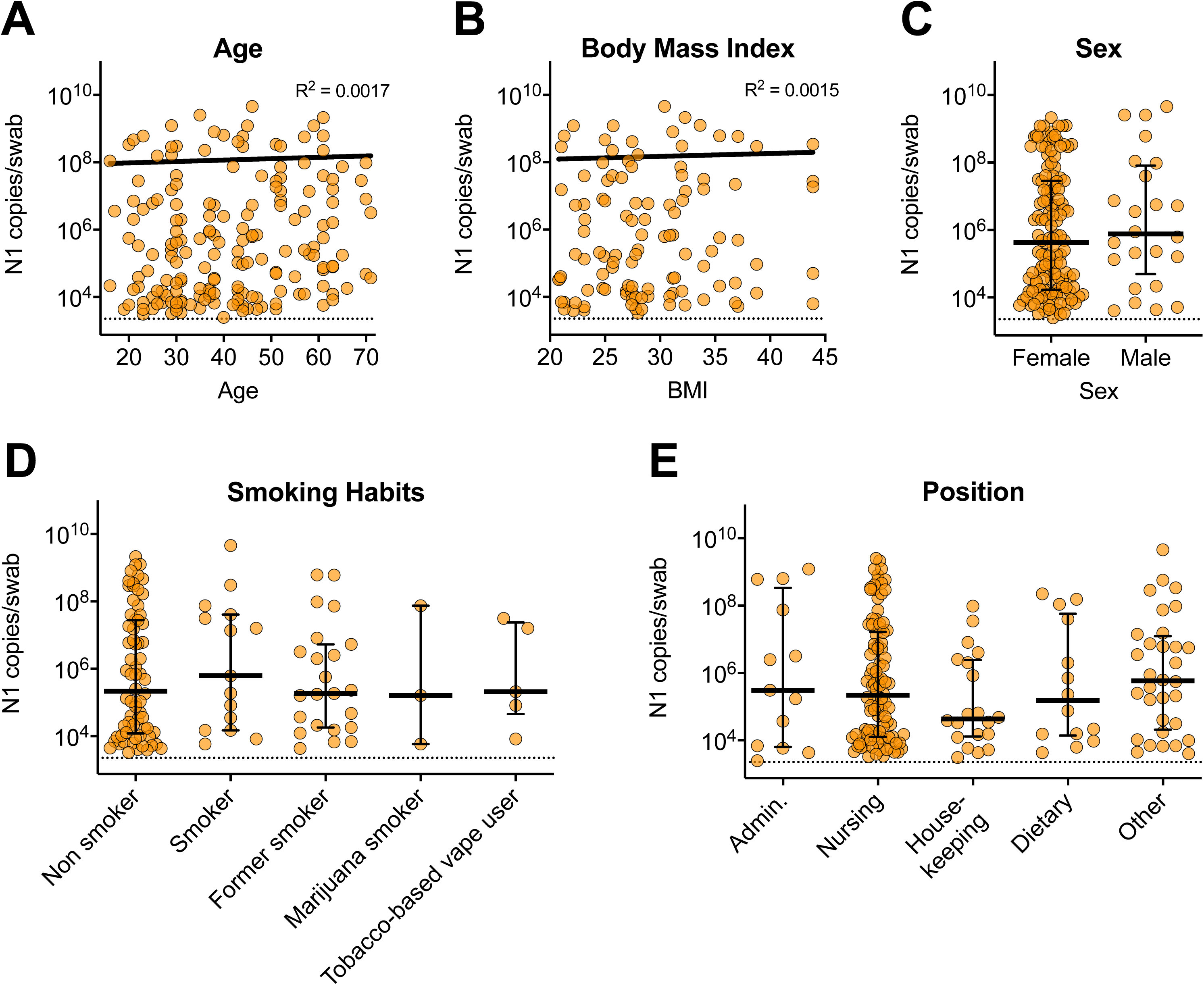
Virus levels stratified by participant age, body mass index, sex and job code. Participants were stratified by **A)** age (n = 91), **B)** BMI (n = 51), **C)** sex (n = 79), **D)** smoking habits, and **E)** job code (n = 90). N1 vRNA from all N1-positive samples were plotted. **A and B)** Semilog nonlinear regression line fit, and **C-D)** bar and errors represent median with interquartile range. Dashed line represents limit of detection.

**Supplemental Figure 2.**
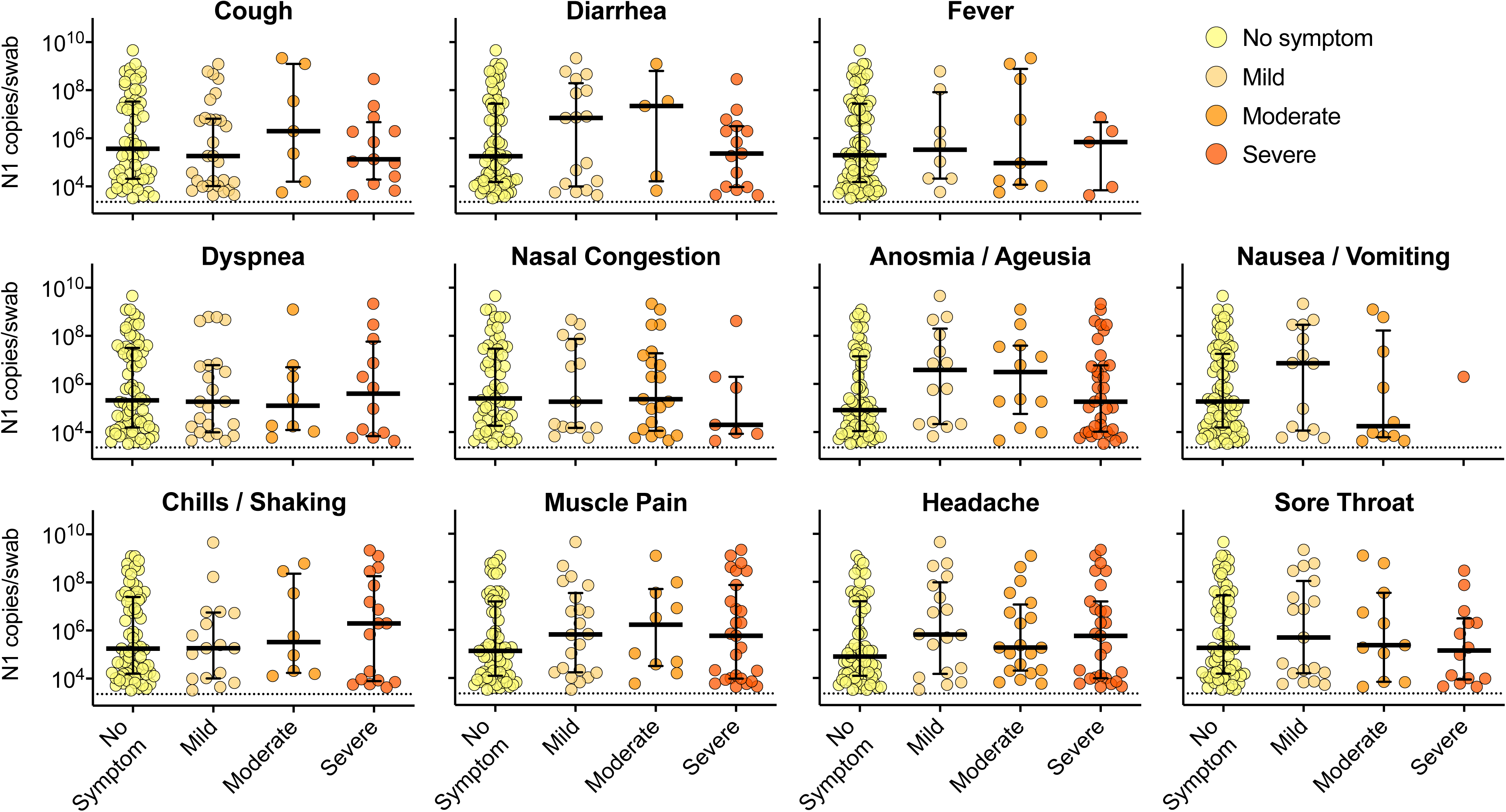
N1 vRNA and symptom severity. N1vRNA levels for each symptom stratified by symptom severity. Bar and errors represent median with interquartile range. Dashed line represents limit of detection.

## Notes

### Competing Interest Statement

The authors have declared no competing interest.

### Author Declarations

This study was reviewed and approved by the Colorado State University IRB under protocol number 20-10057H. Participants provided consent to participate in the study and were promptly informed of test results.

### Summary of Updates

This version is updated with additional data on subject symptoms and demographics. The sequences have been modified to remove repeat samples from the same individuals and new subject sequences are also now included.

